# Estimating the effects of accelerated partner therapy (APT) on reinfection of index cases with chlamydia: a mathematical modelling study

**DOI:** 10.1101/2021.08.11.21261692

**Authors:** Christian L. Althaus, Claudia S. Estcourt, Nicola Low

## Abstract

**Objectives:** The expected effects of accelerated partner therapy (APT) on the reinfection of treated index cases by untreated partners with *Chlamydia trachomatis* (chlamydia) remain unclear. We analyzed data from the LUSTRUM cluster cross-over randomised controlled trial (RCT) using a mathematical model and quantified the effects of offering APT on the probability of successful partner treatment.

**Methods:** We extended a previously developed mathematical model to compute the probability of chlamydia reinfection of index cases by their untreated partners with chlamydia. We fitted the model to data from the RCT and estimated the probability of successful treatment of the partner of index cases using a Bayesian framework.

**Results:** We estimated the median probability of reinfection with chlamydia at 16.3% (50% credible interval, CrI: 12.7-20.0%) without partner treatment and 2.3% (50% CrI: 1.7-3.6%) when partner treatment is 100% successful. The observed rates of reinfection in the RCT were 6.7% (95% confidence interval, CI: 5.6-8.0%) during the control period (standard partner notification (PN)) and 4.8% (95% CI: 3.7-5.9%) during the intervention period where APT was offered in addition to standard PN. These rates correspond to a median probability of successful partner treatment of 63% (50% CrI: 47-76%) during the control period and 78% (50% CrI: 65-87%) during the intervention period.

**Conclusions:** Our study suggests that the observed reduction in reinfection with chlamydia when offering APT is consistent with a higher probability of successful partner treatment.

## INTRODUCTION

Reinfection of treated index cases by untreated partners with *Chlamydia trachomatis* (chlamydia) is common and can lead to further transmission and increases the risk of complications after infection.^1^ Partner notification (PN), the process of identifying, testing and treating sex partners of index cases with a sexually transmitted infection (STI), plays a key role in limiting reinfection in people who have been treated for chlamydia.^2^ Improved PN methods can offer the opportunity to prevent reinfection of index cases more effectively. Expedited partner therapy (EPT) is a method of enhanced patient referral where sexual partners receive medication from the index cases without prior medical evaluation.^1^ Accelerated partner therapy (APT) is an adaptation of EPT and additionally includes a clinical assessment of sex partners through telephone-led or face-to-face consultation with an appropriately qualified healthcare professional.^3^ Since APT aims to reduce the time to partner treatment and increase the proportion of partners treated, it is important to quantify the potential effects of APT on the reinfection of index cases with chlamydia.

The Limiting Undetected Sexually Transmitted infections to RedUce Morbidity (LUSTRUM) cluster cross-over randomised controlled trial (RCT) in the UK aims to determine the effectiveness of APT among opposite-sex partners with chlamydia.^4^ The primary outcome of the RCT was the proportion of index patients with a positive chlamydia test 12-24 weeks after treatment.^5^ The LUSTRUM RCT is accompanied by a dynamic transmission model of chlamydia to study the potential effects of APT at the population level and a cost-effectiveness analysis of APT compared to standard PN over a period of 5 years.^6 7^ Mathematical models can also be used to study the effects of different PN strategies on the reinfection of index cases with chlamydia and thus offer a helpful tool to better understand the results from the RCT.^2^

In this study, we analyzed the outcome of the LUSTRUM cluster cross-over randomised controlled trial (RCT) using a mathematical model and quantified the effects of offering APT in addition to standard PN on the probability of successful partner treatment. We estimated the probability of successful treatment of the partner of index cases after the control and intervention period of the RCT.

## METHODS

### Mathematical model

We extended a previously published mathematical model to compute the probability of chlamydia reinfection of index cases by their untreated sexual partners:^2^

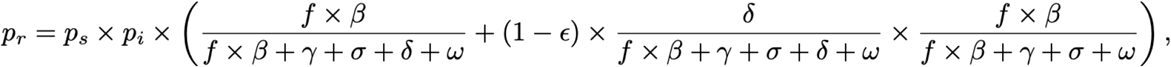

where *p*_*s*_ is the probability that the index case will have future sex with the partner and *p*_*i*_ is the chlamydia-positivity in the infected partner. The first term in the brackets corresponds to the probability that a successfully treated index case is reinfected before the infection in the partner is cleared, the sexual partnership ends, the partner is successfully treated, or the follow-up period ends. *f* denotes the frequency of sex acts and *β* is the per sex act transmission probability between the index case and partner. 1/*γ* is the average duration of chlamydia infection in the infected partner and 1/*σ* is the average duration of the sexual partnership. 1/*δ* is the average time to partner treatment and 1/*ω* corresponds to the follow-up period. The second term in the brackets represents the probability of reinfection of index cases when the partner has not been successfully treated. *ε* is the probability of successful treatment of the partner of index cases.

### Parameters and Bayesian inference

We first calculated the expected probability of chlamydia reinfection *p*_*r*_ as a function of the probability of successful partner treatment. To this end, we sampled 10^5^ parameter sets from uniform (*U*(*a, b*) from *a* to *b*) or binomial (*B*(*n, p*) with sample size *n* and probability *p*) distributions that were informed by the RCT or literature. *p*_*s*_ was drawn from *B*(2589, 47%), which corresponds to the reported probability that the index case will have future sex with the partner in the control arm of the RCT.^5^ *p*_*i*_ was drawn from the overall chlamydia-positivity in partners who returned the STI self-sampling kit in the RCT (*B*(120, 65%)).^5^ The frequency of sex acts *f* and the per sex act transmission probability *β* were drawn from *U*(1, 7) per day and *U*(6%, 16.7%), respectively.^2^ The average duration of infection 1/*γ* and sexual partnership 1/*σ* were drawn from *U*(6, 12) months and *U*(1 week, 6 months).^2^ The average time to partner treatment 1/*δ* was set to 3.2 days.^6 8^ Finally, the follow-up period for the primary outcome of chlamydia-positivity was drawn from *U*(12, 24) weeks.^5^

In a second step, we generated prior distributions of the expected probability of chlamydia reinfection during the control and intervention period of the RCT. We used *p*_*s*_ for the control period as described above and drew from *B*(2218, 52%) for the intervention period.^5^ Furthermore, we drew the probability of successful treatment of the partner *ε* from *U*(0, 1). We then used Approximate Bayesian Computation to generate posterior distributions of *ε* for the control and intervention period by rejecting those parameter sets where *p*_*r*_ was outside the 95% confidence interval (CI) of the chlamydia-positivity in index cases after the control and intervention period, i.e., the primary outcome of the RCT. The reported chlamydia-positivity from the RCT was 6.7% (95% CI: 5.6-8.0%) after the control period and 4.8% (95% CI: 3.7-5.9%) after the intervention period where APT was offered in addition to standard PN.^5^

## RESULTS

Increasing the probability of successful partner treatment can substantially reduce the probability of reinfection in the modelled population of the RCT (figure 1). Without partner treatment, the median probability of reinfection with chlamydia would be 16.3% (50% credible interval, CrI: 12.7-20.0%) after the follow-up period of 12-24 weeks. The British Association for Sexual Health and HIV (BASHH) recommends at least 0.6 contacts per index case for effective PN for chlamydia.^9^ Assuming 60% successful partner treatment in the modelled RCT, we estimated the probability of reinfection of the index case at 7.9% (50% CrI: 6.1-10.1%). Finally, the estimated probability of reinfection drops to 2.3% (50% CrI: 1.7-3.6%) for the case when partner treatment is 100% successful. In general, every 10% increase in the probability of successful partner treatment results in 1.4% (95% CrI: 0.4-2.1%) absolute reduction in the probability of reinfection.

**Figure 1.**
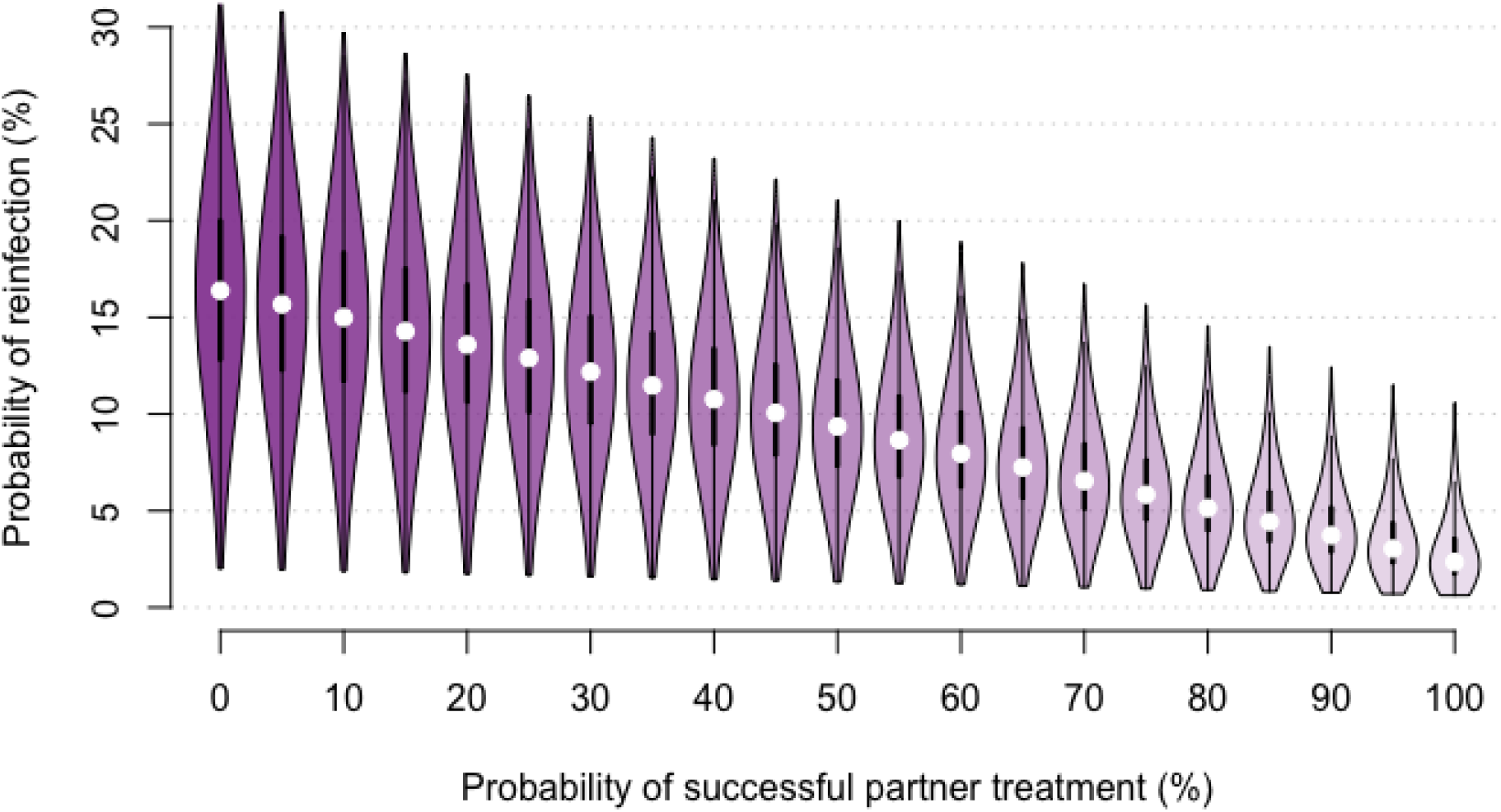
Probability of chlamydia reinfection of index cases by their partners. Posterior distributions of the probability of reinfection are based on 10^5^ randomly sampled parameter sets.

We estimated the probability of successful treatment of the partner of index cases during the control and intervention period of the RCT. We found that the observed chlamydia-positivity after the control period is consistent with a median probability of successful partner treatment of 63% (50% CrI: 47-76%). The lower chlamydia-positivity after the intervention period requires a higher probability of successful partner treatment of 78% (50% CrI: 65-87%). Hence, the model suggests that offering APT in addition to standard PN in the intervention period increased the probability of successful partner treatment by 14% (50% CrI: -4-32%).

## DISCUSSION

We used a mathematical model to study the effects of PN on reinfection of index cases with chlamydia and to analyze the primary outcome of the LUSTRUM cluster cross-over RCT. We showed that the probability of reinfection is expected to decrease linearly with increasing partner treatment. We estimated that offering APT in addition to standard PN in the intervention period of the RCT increased the probability of successful partner treatment from 63% (50% CrI: 47-76%) during the control period to 78% (50% CrI: 65-87%) during the intervention period.

The strength of our study is the application of a mathematical model to study the outcome of an RCT. Using a Bayesian framework allowed us to take into account the considerable uncertainty in critical model parameters, such as the per sex act transmission probabiliy of chlamydia or the average duration of the sexual partnership between the index case and the partner. Furthermore, we were able to include additional data from the RCT in our model, such as the likelihood of future sex with a partner or the chlamydia-positvity in partners who returned the STI self-sampling kit. Nevertheless, there are a number of limitations. First, we did not consider reinfection in women and men separately due to the limited sample size of the RCT. We expect the benefits of offering APT to women and men to be similar, however. Second, the presented mathematical model considers reinfection and treatment from the current partner only. Third, due to the relatively small difference in the primary outcome of the RCT and the use of broad prior parameter distributions, the estimated increase in the probability of successful partner treatment during the intervention period comes with uncertainty.

Reinfection of index cases with chlamydia as estimated by the model and observed in the RCT was lower than findings from an earlier RCT on the effects of expedited partner therapy (EPT) that showed persistent or recurrent chlamydia infection in 13% of participants assigned to standard PN and 10% assigned to EPT.^1^ The reinfection rates were also considerably lower than the observed 15% positivity of repeated tests following a positive chlamydia test in an analysis of health insurance claims data.^10^ However, the observed rates of reinfection in the LUSTRUM cluster cross-over RCT are consistent with the modelled estimates and resulted in an estimated probability of successful partner treatment after the control period of 63%. This estimate is consistent with the BASHH recommendation to contact at least 0.6 partners of index cases for effective PN.^9^ The higher probability of successful partner treatment of 78% during the intervention period is also consistent with a) the higher proportions of more than one sex partner treated and b) the higher proportion of treated partners as observed in the RCT.^5^

This study helps to better understand the outcomes of the LUSTRUM cluster cross-over RCT. Our analysis illustrates that the reported chlamydia-positivity in index cases after the intervention period compared to the control period is consistent with a higher probability of successful partner treatment. Hence, our results suggest that offering APT in addition to standard PN increases the probability of successful partner treatment and thereby reduces reinfection of index cases with chlamydia.

## Data Availability

NA

## Contributors

All authors conceived and designed the study. CLA developed the mathematical model, conducted the analysis, and wrote the manuscript. All authors contributed to the interpretation of the results, commented on the manuscript, and approved the final version of the manuscript.

